# Evaluation of albumin kinetics in mechanically ventilated patients with COVID-19 compared to those with sepsis-induced ARDS

**DOI:** 10.1101/2021.03.16.21253405

**Authors:** Chang Su, Katherine Hoffman, Xu Zhenxing, Elizabeth Sanchez, Ilias Siempos, John S Harrington, Alexandra Racanelli, Maria Plataki, Fei Wang, Edward J. Schenck

## Abstract

COVID-19 outcomes like mortality have been associated with albumin alteration. However, it is unclear whether albumin changes in COVID-19 are pathogen specific or not. To this end, we characterized the kinetics of serum albumin in mechanically ventilated patients with COVID-19 compared to mechanically ventilated patients with sepsis-induced Acute Respiratory Distress Syndrome (ARDS). We discovered two phases of alterations in albumin levels during the course of Covid-19 critical illness, but not for the sepsis-induced ARDS. Our findings suggest the metabolic effects of COVID-19 are pathogen-specific and albumin recovery may signal the cessation of a deleterious immune response in this disease.

## [Main text]

COVID-19 has been associated with alterations in cellular metabolism, including many acute phase proteins such as albumin.^1^ Indeed, a lower albumin at admission to the hospital has been associated with a higher mortality in COVID-19.^2^ However, it is unclear whether albumin changes in COVID-19 are pathogen specific or are due to interventions such as mechanical ventilation. Our objective was to characterize the kinetics of serum albumin in mechanically ventilated patients with COVID-19 compared to mechanically ventilated patients with sepsis-induced Acute Respiratory Distress Syndrome (ARDS).

## Methods

We did a retrospective study at the New York Presbyterian Hospital-Weill Cornell Medical Center (NYP-WCMC) that compared two critically ill cohorts with COVID-19 ^3^ and sepsis-induced ARDS ^4^, respectively. Adult patients in the COVID-19 cohort were admitted from Mar 3 to Jul 10, 2020. SARS-CoV2 diagnosis was made through reverse-transcriptase–PCR assays performed on nasopharyngeal swabs. The critical care response to the pandemic has been previously described ^5^. Adult patients with sepsis-induced ARDS, defined by Sepsis III criteria ^6^ and PaO_2_/FiO_2_ ratio < 300 while treated with mechanical ventilation, were admitted from Dec 12, 2006 to Feb 26, 2019.

We evaluated serum albumin levels from 7 days prior to 30 days after intubation in each cohort. We averaged albumin values over 24 hours if more than 1 value was available. We then examined the albumin progression trajectories, aligned at intubation time to test the relationship at a similar point in disease progression, in survivors and non-survivors. We hypothesized that albumin recovery would be apparent in COVID-19 survivors. We derived an algorithm based on Chow’s test ^7^ to detect the albumin trajectory breakpoint for each patient, where a deteriorating albumin trend changed to a recovering trend. Specifically, for each patient, the Chow’s test was performed for each time point of the albumin trajectory, and the breakpoint was determined by rejection (*P* < 0.05 and F-value ≥ 3) of the null hypothesis that the coefficients of linear regressions before and after the breakpoint are equal. After that, we fit linear mixed-effects models to estimate the deteriorating and recovering trajectories, for survivors and non-survivors, respectively, adjusting for age and sex. We then analyzed albumin trajectories within the sepsis-induced ARDS cohort for comparison.

We reported descriptive data as mean (standard deviation [SD]) or median (interquartile range [IQR]) for continuous variables and number (percentage) for categorical variables. We assessed the differences between groups using Fisher’s exact test for categorical values, and two-sample t test or Wilcoxon rank-sum test for continuous values where appropriate. All the tests were two-sided with a significance level of 0.05.

## Results

The COVID-19 cohort consists of 308 intubated patients with confirmed SARS-CoV2 infection (age 61.7 [SD=16.4] years; 31.5% female). 102 COVID-19 patients died prior to extubation during their hospitalization and 206 were successfully extubated. The non-survivors were older than the survivors (67.3 [SD=12.1] years vs 58.9 [SD=14.9] years, *P*<0.001). There was no significant difference of comorbidities between the non-survivors and survivors. The albumin level 24 h before intubation was similar in survivors and non-survivors (2.42 [SD=0.56] g/dL vs 2.57 [SD=0.58] g/dL, *P*=0.095), while the albumin value 24 h after intubation was lower in non-survivors than the survivors (2.03 [SD=0.50] g/dL vs 2.18 [SD=0.44] g/dL, *P*=0.015).

The sepsis-induced ARDS cohort contains 363 intubated patients with confirmed sepsis (age 68.8 [SD=17.3] years; 40.8% female), of which 52 were non-survivors and 311 were survivors. Overall, the intubated patients with sepsis-induced ARDS showed a higher burden of chronic comorbidities than the COVID-19 patients. Compared to the sepsis-induced ARDS patients, intubated COVID-19 patients had a longer intubation duration (for non-survivors, 18.1 [SD=15.5] days vs 12.8 [SD=17.5] days, *P*=0.070; for survivors, 24.5 [SD=17.8] days vs 8.3 [SD=12] days, *P*<0.001) and higher baseline Sequential Organ Failure Assessment score (for non-survivors, 13 [IQR, 11-15] vs 11 [IQR, 9-14], *P*=0.002; for survivors, 12 [IQR, 11-13] vs 9 [IQR, 7-11], *P*<0.001). More details of the characteristics of the two cohorts were shown in Table 1.

**Table 1.** Clinical characteristics of the studied COVID-19 and sepsis cohorts.

| Variable | COVID-19 cohort |  |  |  | Sepsis cohort |  |  |  |
| --- | --- | --- | --- | --- | --- | --- | --- | --- |
|  | Total | Non-survivors | Survivors | P Value<sup>a</sup> | Total | Non-survivors | Survivors | P Value<sup>a</sup> |
| Number of patients | 308 | 102 | 206 | - | 363 | 52 | 311 | - |
| Demographics |  |  |  |  |  |  |  |  |
| Age, years, Mean (SD) | 61.7 (14.6) | 67.3 (12.1) | 58.9 (14.9) | <0.001 | 68.8 (17.3) | 72.2 (17.5) | 68.3 (17.2) | 0.140 |
| Sex female, n (%) | 97 (31.5) | 27 (26.5) | 70 (34.0) | 0.194 | 148 (40.8) | 16 (30.8) | 132 (42.4) | 0.128 |
| Race white, n (%) | 96 (31.2) | 33 (32.4) | 63 (30.6) | 0.794 | 116 (32.0) | 17 (32.7) | 99 (31.8) | 0.874 |
| Body mass index, kg/m<sup>2</sup>, Mean (SD) | 29.3 (8.1) | 28.6 (7.8) | 29.7 (8.2) | 0.183 | 29.1 (12.7) | 27.3 (6.7) | 29.4 (13.4) | 0.195 |
| Comorbidities, n (%) |  |  |  |  |  |  |  |  |
| Active Cancer (liquid) | 12 (3.9) | 7 (6.9) | 5 (2.4) | 0.067 | 32 (8.8) | 12 (23.1) | 20 (6.4) | <0.001 |
| Active Cancer (solid) | 8 (2.6) | 4 (3.9) | 4 (1.9) | 0.447 | 30 (8.3) | 3 (5.8) | 27 (8.7) | 0.596 |
| Congestive heart failure | 57 (18.5) | 22 (21.6) | 35 (17.0) | 0.351 | 131 (36.1) | 18 (34.6) | 113 (36.3) | 0.877 |
| Hypertension | 166 (53.9) | 59 (57.8) | 107 (51.9) | 0.334 | 209 (57.6) | 25 (48.1) | 184 (59.2) | 0.172 |
| Pulmonary Disease | 60 (19.5) | 23 (22.5) | 37 (18.0) | 0.360 | 103 (28.4) | 15 (28.8) | 88 (28.3) | 1 |
| Diabetes Mellitus | 93 (30.2) | 33 (32.4) | 60 (29.1) | 0.598 | 93 (25.6) | 10 (19.2) | 83 (26.7) | 0.305 |
| Renal Disease | 24 (7.8) | 10 (9.8) | 14 (6.8) | 0.371 | 91 (25.1) | 16 (30.8) | 75 (24.1) | 0.304 |
| Liver Disease | 6 (1.9) | 4 (3.9) | 2 (1.0) | 0.095 | 63 (17.4) | 15 (28.8) | 48 (15.4) | 0.028 |
| Duration of mechanical ventilation, days, Mean (SD) | 22.4 (17.3) | 18.1 (15.5)<sup>b</sup> | 24.5 (17.8)<sup>c</sup> | 0.002 | 8.9 (13.0) | 12.8 (17.5)<sup>b</sup> | 8.3 (12.0)<sup>c</sup> | 0.031 |
| P/F ratio, within 24 h after intubation, Mean (SD) | 167.6 (90.2) | 166.6 (106.6)<sup>d</sup> | 168.1 (80.9)<sup>e</sup> | 0.612 | 192.9 (141.0) | 139.4 (84.8)<sup>d</sup> | 201.8 (146.4)<sup>e</sup> | <0.001 |
| SOFA, within 24 h after intubation, Median (IQR) | 12 (11-14) | 13 (11-15)<sup>f</sup> | 12 (11-13)<sup>g</sup> | <0.001 | 9.0 (7 - 11) | 11 (9 - 14)<sup>f</sup> | 9 (7 - 11)<sup>g</sup> | <0.001 |
| Albumin level, g/dL, Mean (SD) |  |  |  |  |  |  |  |  |
| Within 24 h before intubation | 2.53 (0.58) | 2.42 (0.56) | 2.57 (0.58) | 0.095 | 2.65 (0.82) | 2.27 (1.00) | 2.72 (0.77) | 0.102 |
| Within 24 h after intubation | 2.14 (0.49) | 2.03 (0.50) | 2.18 (0.44) | 0.015 | 2.37 (0.58) | 2.17 (0.58) | 2.41 (0.57) | 0.04 |
| Breakpoint Albumin levels |  |  |  |  |  |  |  |  |
| Time to Breakpoint, days (after intubation), Mean (SD) | 5.50 (3.88) | 5.61 (4.06) | 5.47 (3.83) | 0.943 | - | - | - | - |
| Albumin level at breakpoint, g/dL, Mean (SD) | 1.61 (0.38) | 1.50 (0.43) | 1.64 (0.36) | 0.030 | - | - | - | - |
| Infection source, n (%) |  |  |  |  |  |  |  |  |
| Pneumonia | - | - | - | - | 157 (43.3) | 29 (55.8) | 128 (41.2) | 0.068 |
| Urinary Tract | - | - | - | - | 71 (19.6) | 4 (7.7) | 67 (21.5) | 0.022 |
| Intra-Abdominal | - | - | - | - | 66 (18.2) | 16 (30.8) | 50 (16.1) | 0.018 |

Definition of abbreviations: IQR = interquartile range; P/F ratio = ratio of arterial oxygen partial pressure (PaO<sub>2</sub>) to fractional inspired oxygen (FiO<sub>2</sub>); SD = standard deviation; SOFA = Sequential Organ Failure Assessment.
<sup>a</sup> The P values were calculated to assess the differences between non-survivors and survivors, using Fisher's exact test for categorical values, and two-sample t test or Wilcoxon rank-sum test for continuous values where appropriate.
<sup>b</sup> $P=0.070$ for cross cohort comparison of intubation duration of non-survivors.
<sup>c</sup> $P<0.001$ for cross cohort comparison of intubation duration of survivors.
<sup>d</sup> $P=0.078$ for cross cohort comparison of baseline P/F ratio of non-survivors.
<sup>e</sup> $P=0.005$ for cross cohort comparison of baseline P/F ratio of survivors.
<sup>f</sup> $P=0.002$ for cross cohort comparison of baseline SOFA score of non-survivors.
<sup>g</sup> $P<0.001$ for cross cohort comparison of baseline SOFA score of survivors.

Albumin trajectory in all critically ill Covid-19 patients consists of two clearly distinct phases (Figure 1). Phase I (deterioration) was defined by rapid albumin loss and Phase II (recovery) showed albumin stabilization or improvement. The Chow’s test detected albumin breakpoint for each patient occurred 5.47 [SD=3.83] days after intubation vs 5.61 [SD=4.06] days after intubation (*P*=0.943), in survivors and non-survivors, while the breakpoint albumin level was lower in non-survivors compared to that in survivors (1.50 [SD=0.43] g/dL vs 1.64 [SD=0.36] g/dL, *P*=0.030) (Table 1 and Figure 1A). Based on the breakpoint for each patient, linear mixed-effects models identified clear deterioration phases with similar slopes (i.e., rates of daily change of albumin level), among non-survivors (β=-0.101, 95% CI, −0.108 to −0.095, *P*<0.001) and survivors (β=-0.104, 95% CI, −0.109 to −0.099, *P*<0.001) (Figure 1A). Following the deterioration phase, there was a recovery phase in survivors (β=0.022, 95% CI, 0.021 to 0.023, *P*<0.001), that was higher than non-survivors (β=-0.002, 95% CI, −0.004 to 0.000, *P*=0.068). In contrast, we did not find a two-phase albumin progression trend in both survivors and non-survivors in the sepsis cohort (Figure 1B).

**Figure 1.**
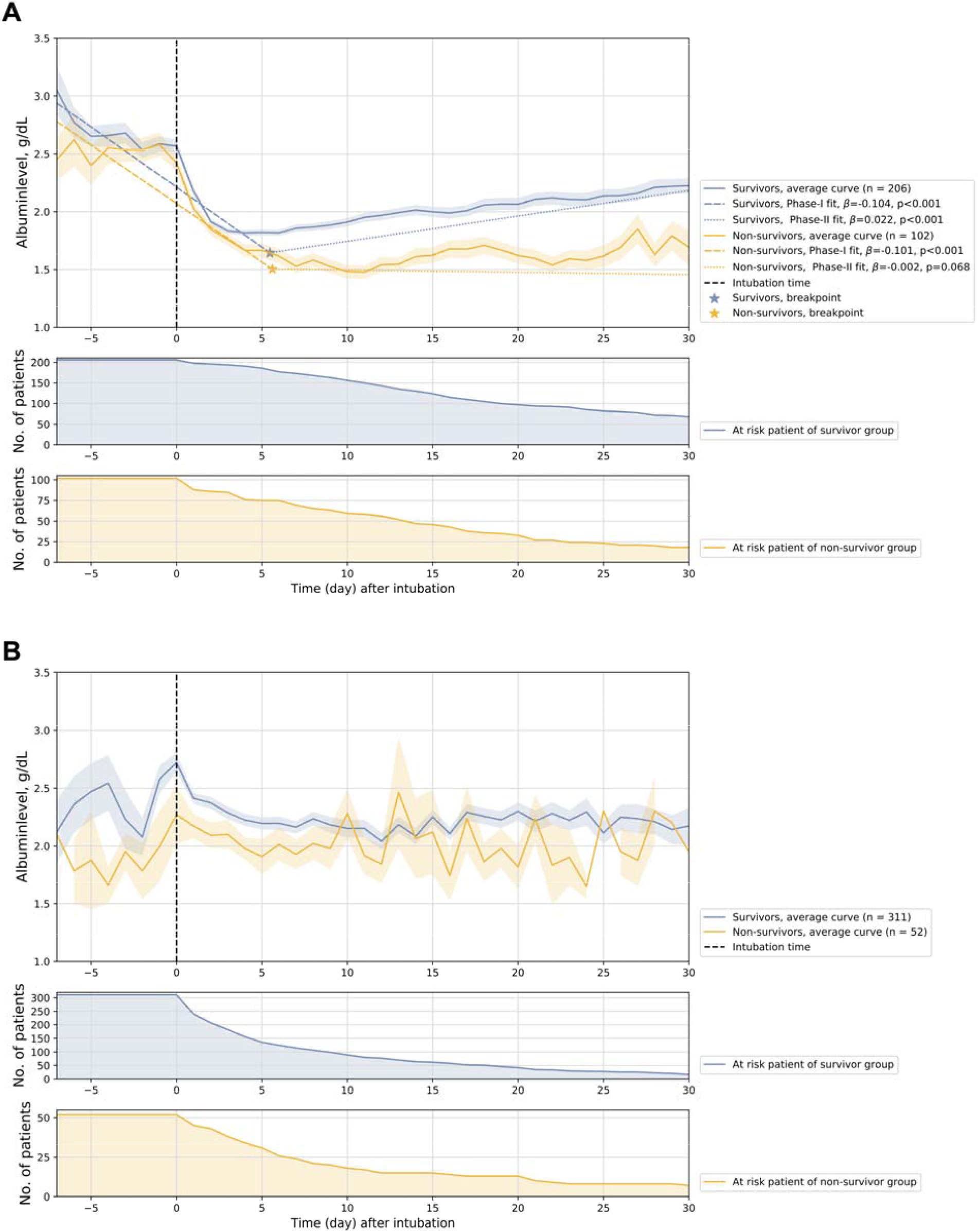
Illustration of albumin trajectories. (A) Albumin trajectories (averaged trajectories and two-phase linear mixed-effects model estimated trajectories) of non-survivors and survivors of the critically ill COVID-19 cohort. (B) Albumin trajectories (averaged) of non-survivors and survivors of the sepsis-induced ARDS cohort.

## Discussion

We defined two phases of alterations in albumin levels during the course of Covid-19 critical illness. Albumin fell rapidly following intubation in our COVID-19 cohort regardless of outcome, however albumin recovery predicted clinical improvement in critical COVID-19. We did not see this pattern in a large sepsis-induced ARDS cohort. In our sepsis-induced ARDS cohort, albumin levels were prognostic; but, we did not see a clear recovering pattern in survivors. The albumin kinetics in sepsis-induced ARDS were similar to the ALBIOS study of albumin resuscitation in sepsis.^8^

Different albumin kinetics in COVID-19 compared to sepsis-induced ARDS may be due to differences in the patient’s premorbid states. Our COVID-19 patients had a lower burden of chronic liver disease, kidney disease and cancer compared to our sepsis population. However, COVID-19 may have temporally distinct metabolic effects compared to other life-threatening infections.^1,9,10^ Lastly, it is unclear whether the lack of albumin recovery in COVID-19 non-survivors was related to ongoing inflammatory viral effects or secondary hospital acquired complications. The metabolic effects of COVID-19 are pathogen-specific and albumin recovery may signal the cessation of a deleterious immune response in this disease.

## Data Availability

Data is not publicly available.

## Author Contributions

EJS, CS and FW conceived of the study. EJS, ES, IIS, AMC, and KH created the data sources and developed case definitions. CS, KH, ZX, FW and EJS did the data analysis. CS and EJS wrote the manuscript and all authors critically edited the final manuscript and approved.

## Ethics approval

Approved by the IRB at WCM #20-04021909, #1811019761

